# Enoxaparin is associated with lower rates of thrombosis, kidney injury, and mortality than Unfractionated Heparin in hospitalized COVID patients

**DOI:** 10.1101/2020.10.06.20208025

**Authors:** Colin Pawlowski, AJ Venkatakrishnan, Christian Kirkup, Gabriela Berner, Arjun Puranik, John C. O’Horo, Andrew D. Badley, Venky Soundararajan

**Affiliations:** nference, inc., One Main Street, Suite 400, East Arcade, Cambridge, MA 02142, USA; Mayo Clinic, Rochester MN, USA

**Author notes:** Address correspondence to VS. Equal contribution (Colin Pawlowski, AJ Venkatakrishnan, Christian Kirkup).

## Abstract

Although anticoagulants such as unfractionated heparin and low molecular weight heparin (LMWH, e.g. enoxaparin) are both being used for therapeutic mitigation of COVID associated coagulopathy (CAC), differences in their clinical outcomes remain to be investigated. Here, we employ automated neural networks supplemented with expert curation (‘augmented curation’) for retrospectively analyzing the complete electronic health records (EHRs) of 671 hospitalized COVID-19 patients administered either enoxaparin or unfractionated heparin, but not both. We find that COVID-19 patients administered unfractionated heparin but not enoxaparin have higher rates of mortality (risk ratio: 2.6; 95% C.I.: [1.2-5.4]; p-value: 0.02; BH adjusted p-value: 0.09), thrombotic events (risk ratio: 5.7, 95% C.I.: [2.1, 33.9], p-value: 0.024), acute kidney injury (risk ratio: 5.5; 95% C.I.: [1.2-17.7]; p-value: 0.02; BH adjusted p-value: 0.10), and bacterial pneumonia (risk ratio undefined; 95% C.I.: [1.0, 292]; p-value:0.02; BH adjusted p-value:0.10), compared to patients administered enoxaparin but not unfractionated heparin. Notably, even after controlling for potential confounding factors such as demographics, comorbidities, admission diagnosis, initial ICU status, and initial level of oxygen support, the above differences between the enoxaparin and unfractionated heparin patient cohorts remain statistically significant. This study emphasizes the need for mechanistically investigating differential modulation of the COVID-associated coagulation cascades by enoxaparin versus unfractionated heparin.

## Introduction

COVID-19 manifests in varying levels of patient outcomes ranging from mild, moderate to severe disease^1–3^. While the mild/asymptomatic patients have been managed through quarantining and self-medication, the optimal management of hospitalized moderately ill or severely ill COVID-19 patients remains a formidable challenge. Owing to the diversity of comorbidities and complications in the afflicted patients and the overwhelming pace of the pandemic the regimen of medications in COVID-19 critical care is yet to be standardized. Meanwhile, the accumulation of real-world data on patient outcomes of COVID-19 from various healthcare systems provides an excellent opportunity to identify underlying trends that could lead to actionable insights.

Coagulopathies are a major class among COVID-19 associated complications^4^, particularly in a critical care setting^5^. Prior studies have provided a fine grained resolution of the hematological parameters in COVID-19 patients^6^. Trials comparing anticoagulation treatments are ongoing, however owing to the wide-spread and severe impact of the disease, there is a need for evaluation of observational data in order to rapidly generate evidence to guide treatment^7^. A spectrum of anticoagulants such as Heparin, Enoxaparin, and Rivaroxaban are being used in COVID-19 patient management as needed^8,9^. A clinical trial is being designed to examine whether prophylactic-dose enoxaparin improves survival and reduces hospitalizations in older (age > 50) symptomatic ambulatory patients^10^. Furthermore, recent in vitro studies have shown that pseudotyped viral particles were efficiently neutralized by a variety of anticoagulants ^11^. In addition, results from a phase II clinical trial found that patients treated with therapeutic enoxaparin had improved gas exchange and more ventilator-free days compared to patients treated with standard anticoagulant thromboprophylaxis^12^. However, the full array of patient outcomes associated with various anticoagulants remains to be understood. The availability of patient outcomes and drugs administered in Mayo Clinic sites and associated health systems enables us to address this question. Here, we present a comparison of the patient outcomes in terms of mortality status, ICU admission and the durations of stay in ICU and hospital in severe COVID-19 patients that were administered Heparin vs. Enoxaparin. We use propensity score matching to construct matched cohorts for comparison which are balanced across a range of clinical covariates, including: demographics, comorbidities, admission diagnosis, initial ICU status, and initial level of oxygen support (see **Methods**). The clinical covariates for the study populations of the original and matched cohorts are presented in **Table 1**. The outcomes for the original and matched cohorts are presented in **Table 2** and **Table 3**, respectively.

**Table 1:**
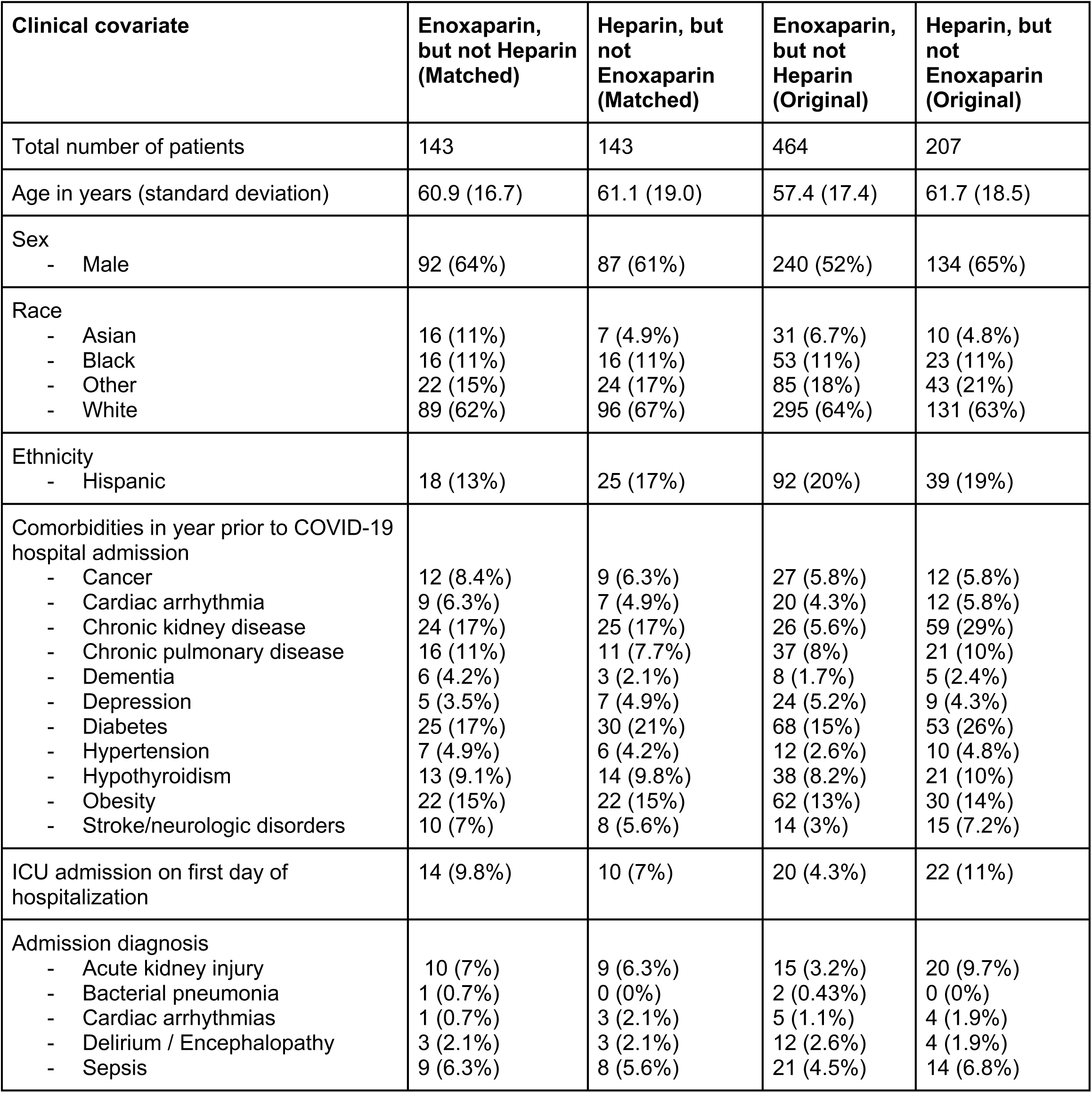
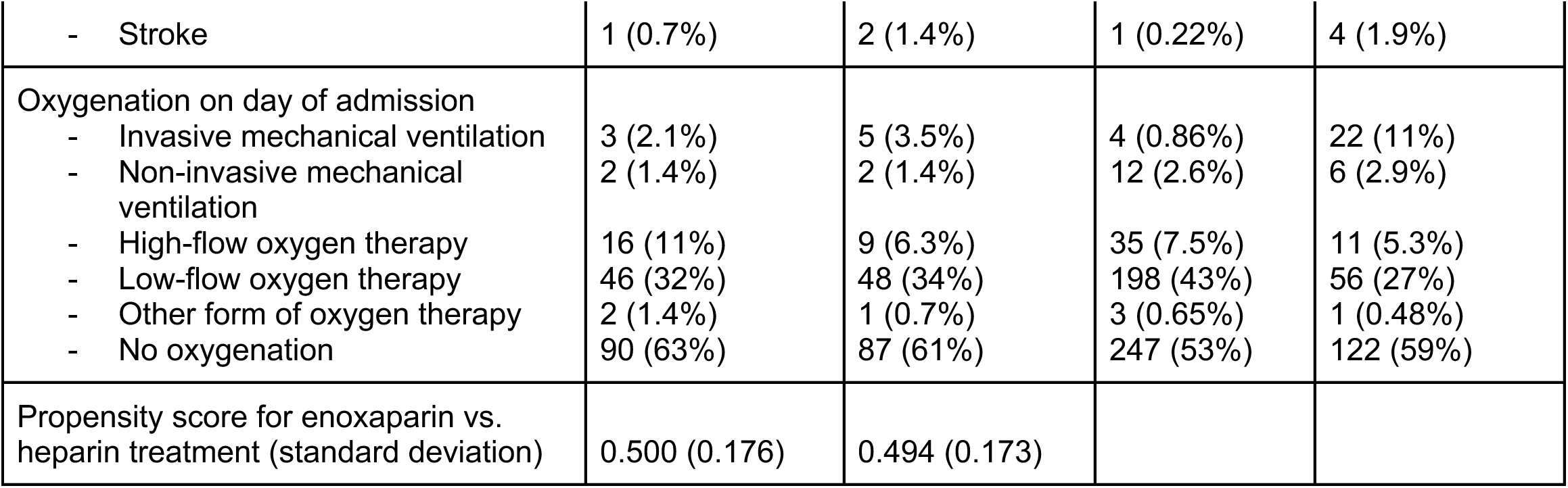
Summary of patient characteristics for matched and original cohorts of hospitalized COVID-19 patients who have taken either heparin or enoxaparin. For numeric variables such as age, the mean value for each cohort is shown with standard deviation in parentheses. For categorical variables such as race and ethnicity, patient counts are shown with the percentage of each cohort in parentheses.

| Clinical covariate | Enoxaparin, but not Heparin (Matched) | Heparin, but not Enoxaparin (Matched) | Enoxaparin, but not Heparin (Original) | Heparin, but not Enoxaparin (Original) |
| --- | --- | --- | --- | --- |
| Total number of patients | 143 | 143 | 464 | 207 |
| Age in years (standard deviation) | 60.9 (16.7) | 61.1 (19.0) | 57.4 (17.4) | 61.7 (18.5) |
| Sex |  |  |  |  |
| - Male | 92 (64%) | 87 (61%) | 240 (52%) | 134 (65%) |
| Race |  |  |  |  |
| - Asian | 16 (11%) | 7 (4.9%) | 31 (6.7%) | 10 (4.8%) |
| - Black | 16 (11%) | 16 (11%) | 53 (11%) | 23 (11%) |
| - Other | 22 (15%) | 24 (17%) | 85 (18%) | 43 (21%) |
| - White | 89 (62%) | 96 (67%) | 295 (64%) | 131 (63%) |
| Ethnicity |  |  |  |  |
| - Hispanic | 18 (13%) | 25 (17%) | 92 (20%) | 39 (19%) |
| Comorbidities in year prior to COVID-19 hospital admission |  |  |  |  |
| - Cancer | 12 (8.4%) | 9 (6.3%) | 27 (5.8%) | 12 (5.8%) |
| - Cardiac arrhythmia | 9 (6.3%) | 7 (4.9%) | 20 (4.3%) | 12 (5.8%) |
| - Chronic kidney disease | 24 (17%) | 25 (17%) | 26 (5.6%) | 59 (29%) |
| - Chronic pulmonary disease | 16 (11%) | 11 (7.7%) | 37 (8%) | 21 (10%) |
| - Dementia | 6 (4.2%) | 3 (2.1%) | 8 (1.7%) | 5 (2.4%) |
| - Depression | 5 (3.5%) | 7 (4.9%) | 24 (5.2%) | 9 (4.3%) |
| - Diabetes | 25 (17%) | 30 (21%) | 68 (15%) | 53 (26%) |
| - Hypertension | 7 (4.9%) | 6 (4.2%) | 12 (2.6%) | 10 (4.8%) |
| - Hypothyroidism | 13 (9.1%) | 14 (9.8%) | 38 (8.2%) | 21 (10%) |
| - Obesity | 22 (15%) | 22 (15%) | 62 (13%) | 30 (14%) |
| - Stroke/neurologic disorders | 10 (7%) | 8 (5.6%) | 14 (3%) | 15 (7.2%) |
| ICU admission on first day of hospitalization | 14 (9.8%) | 10 (7%) | 20 (4.3%) | 22 (11%) |
| Admission diagnosis |  |  |  |  |
| - Acute kidney injury | 10 (7%) | 9 (6.3%) | 15 (3.2%) | 20 (9.7%) |
| - Bacterial pneumonia | 1 (0.7%) | 0 (0%) | 2 (0.43%) | 0 (0%) |
| - Cardiac arrhythmias | 1 (0.7%) | 3 (2.1%) | 5 (1.1%) | 4 (1.9%) |
| - Delirium / Encephalopathy | 3 (2.1%) | 3 (2.1%) | 12 (2.6%) | 4 (1.9%) |
| - Sepsis | 9 (6.3%) | 8 (5.6%) | 21 (4.5%) | 14 (6.8%) |
| - Stroke | 1 (0.7%) | 2 (1.4%) | 1 (0.22%) | 4 (1.9%) |
| Oxygenation on day of admission |  |  |  |  |
| - Invasive mechanical ventilation | 3 (2.1%) | 5 (3.5%) | 4 (0.86%) | 22 (11%) |
| - Non-invasive mechanical ventilation | 2 (1.4%) | 2 (1.4%) | 12 (2.6%) | 6 (2.9%) |
| - High-flow oxygen therapy | 16 (11%) | 9 (6.3%) | 35 (7.5%) | 11 (5.3%) |
| - Low-flow oxygen therapy | 46 (32%) | 48 (34%) | 198 (43%) | 56 (27%) |
| - Other form of oxygen therapy | 2 (1.4%) | 1 (0.7%) | 3 (0.65%) | 1 (0.48%) |
| - No oxygenation | 90 (63%) | 87 (61%) | 247 (53%) | 122 (59%) |
| Propensity score for enoxaparin vs. heparin treatment (standard deviation) | 0.500 (0.176) | 0.494 (0.173) |  |  |

**Table 2:** Summary of clinical outcomes for **unmatched** cohorts of hospitalized COVID-19 patients who have taken either heparin or enoxaparin. For categorical variables such as mortality status and complications, patient counts are shown with the percentage of each cohort in parentheses. For numeric variables such as hospital and ICU length of stay, the mean value for each cohort is shown with standard deviation in parentheses. In addition, Benjamini-Hochberg adjusted p-values are shown for the statistical tests comparing the outcome variables for the matched Enoxaparin and Heparin cohorts.

| Outcome variable | Enoxaparin, but not Heparin (Original) | Heparin, but not Enoxaparin (Original) | Chi-square p-value | BH-adjusted p-value |
| --- | --- | --- | --- | --- |
| Number of patients | 464 | 207 |  |  |
| Deceased (ever) | 14 (3%) | 32 (15%) | 1.0e-8 | 1.7e-8 |
| Deceased 28 days within first day of hospitalization (out of patients with known mortality status) | 12 (4.7%) | 23 (15%) | 4.4e-4 | 4.4e-4 |
| ICU admission | 90 (19%) | 70 (34%) | 7.8e-5 | 9.8e-5 |
| Hospital length of stay in days | 5.4 (4.2) | 8.0 (7.2) | 4.6e-9 | 1.5e-8 |
| ICU length of stay in days | 0.90 (2.5) | 2.8 (6.0) | 6.1e-9 | 1.5e-8 |

**Table 3:** Summary of clinical outcomes for **matched** cohorts of hospitalized COVID-19 patients who have taken either heparin or enoxaparin. For categorical variables such as mortality status and complications, patient counts are shown with the percentage of each cohort in parentheses. For numeric variables such as hospital and ICU length of stay, the mean value for each cohort is shown with standard deviation in parentheses. In addition, Benjamini-Hochberg adjusted p-values are shown for the statistical tests comparing the outcome variables for the matched Enoxaparin and Heparin cohorts.

| Outcome variable | Enoxaparin, but not Heparin (Matched) | Heparin, but not Enoxaparin (Matched) | Chi-square p-value | BH-adjusted p-value |
| --- | --- | --- | --- | --- |
| Number of patients | 143 | 143 |  |  |
| Deceased (ever) | 8 (5.6%) | 21 (15%) | 0.02 | 0.09 |
| Deceased 28 days within first day of hospitalization (out of patients with known mortality status) | 6 (7.7%) | 15 (16%) | 0.16 | 0.21 |
| ICU admission | 37 (26%) | 41 (29%) | 0.69 | 0.69 |
| Hospital length of stay in days | 5.8 (4.9) | 6.9 (6.3) | 0.10 | 0.17 |
| ICU length of stay in days | 1.1 (2.4) | 1.9 (4.7) | 0.10 | 0.17 |

## Results

### Patients that were administered enoxaparin have lower mortality rates, lower ICU admission rates, and shorter hospital / ICU stays

We compared the mortality rate and ICU admission rate in patients from the curated Mayo Clinic dataset of 671 hospitalized COVID-19 patients who received either enoxaparin or heparin, but not both. Of these patients, 464 were administered enoxaparin but not unfractionated heparin; of these 14 (3%) were deceased. For comparison, 207 patients were administered heparin but not enoxaparin. Of these patients, 32 (15%) were deceased. Comparing the mortality outcomes, patients in the heparin only cohort have a higher mortality rate than those in the opposing enoxaparin cohort (risk ratio of death: 5.12; 95% C.I.: [2.76, 9.10]; adjusted p-value 1.7e-8) (**Table 2**). Of the 464 patients administered enoxaparin but not heparin, 90 (19%) were later admitted to the ICU. Similarly, of the 207 patients administered heparin but not enoxaparin, 70 (34%) were admitted to the ICU. Comparing the ICU admission status, patients administered heparin had a higher admission to ICU rate compared to Enoxaparin (risk ratio of ICU admission: 1.74; 95% C.I.: [1.34, 2.27]; adjusted p-value 9.8e-5) (**Table 2**).

Next we compared the lengths of stay in the ICU and the overall length of stay in the hospital. Here, we restricted the analysis to only patients that were alive. The length of stay in the ICU and in the hospital were shorter for patients administered enoxaparin but not heparin (mean ICU duration: 0.9 days, mean hospital duration: 5.4 days) compared to the patients administered heparin but not enoxaparin (mean ICU duration: 2.8 days, mean hospital duration: 8.1 days) (**Table 2**). The difference in hospital length of stay and ICU length of stay across the two cohorts are both statistically significant according to a t-test (hospital duration adjusted p-value: 1.5e-8; ICU duration adjusted p-value: 1.5e-8). Taken together, this suggests preliminarily that enoxaparin is correlated with a lower mortality rate, ICU admission rate and ICU and hospital length of stay compared to heparin. Also of note are the increased rates of acute kidney injury (risk ratio: 5.2; 95% C.I.: [2.68-9.71]; adjusted p-value 7.8e-7), bacterial pneumonia (risk ratio: 5.37; 95% C.I.: [1.89-13.67]; adjusted p-value 2.6e-3) and co-/secondary infection (risk ratio: 5.97; 95% C.I.: [2.32-13.86]; adjusted p-value 1.9e-4) in the heparin cohort (**Table 4**). On examination of cause of death for deceased patients for both cohorts, it is clear that the majority of deaths occurring in both the heparin and enoxaparin cohorts are driven primarily by COVID-19 associated causes giving an indication that divergent rates of mortality between the cohorts is not the result of extraneous illness or accidents (**Table 5**).

**Table 4:**
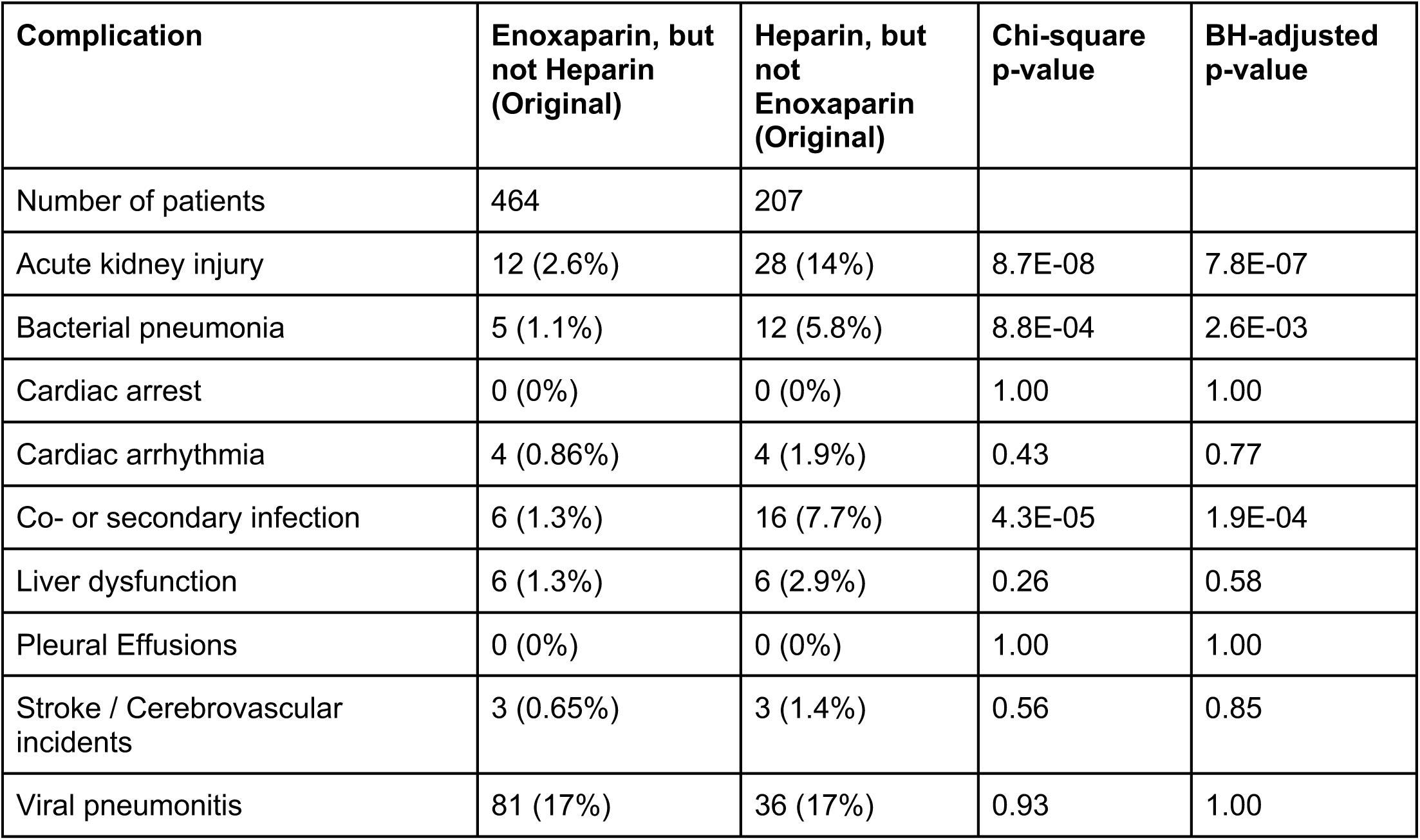
Summary of occurrences of complications during hospitalization (days 0 to 28) for **unmatched** cohorts of hospitalized COVID-19 patients who have taken either heparin or enoxaparin.

**Table 5:** Cause of death information for patients administered enoxaparin only or heparin only. Each row corresponds to a patient, with fields indicating whether the patient had one of the following conditions: ARDS, AKI, Sepsis, Pneumonia, Heart Failure, or Other (any other medical condition). In addition, the last column indicates if the cause of death could not be determined for the particular patient.

| **Anticoagulant** | **ARDS/Acute**<br>**respiratory**<br>**failure/Hypoxia** | **AKI/Renal**<br>**Failure** | **Sepsis** | **Pneumonia** | **Heart**<br>**Failure** | **Other** | **Inconclusive** |
| --- | --- | --- | --- | --- | --- | --- | --- |
| Enoxaparin | yes |  |  | yes |  |  |  |
| Enoxaparin |  | yes |  |  |  | yes |  |
| Enoxaparin |  | yes |  |  |  | yes |  |
| Enoxaparin | yes |  |  | yes |  |  |  |
| Enoxaparin |  |  |  | yes |  | yes |  |
| Enoxaparin |  |  |  |  |  | yes |  |
| Enoxaparin | yes | yes |  |  | yes | yes |  |
| Enoxaparin | yes |  |  | yes |  |  |  |
| Heparin | yes | yes | yes | yes |  |  |  |
| Heparin | yes | yes |  |  | yes |  |  |
| Heparin |  |  |  |  |  |  | yes |
| Heparin |  |  |  |  |  |  | yes |
| Heparin | yes |  | yes | yes |  |  |  |
| Heparin | yes |  | yes | yes |  |  |  |
| Heparin | yes | yes |  | yes |  |  |  |
| Heparin |  |  |  |  |  |  | yes |
| Heparin | yes |  | yes | yes | yes |  |  |
| Heparin | yes | yes |  | yes |  |  |  |
| Heparin | yes |  |  | yes | yes |  |  |
| Heparin | yes | yes |  | yes |  |  |  |
| Heparin | yes |  |  | yes |  |  |  |
| Heparin | yes |  |  | yes |  |  |  |
| Heparin | yes |  | yes | yes |  |  |  |
| Heparin | yes |  |  | yes | yes |  |  |
| Heparin |  |  |  |  |  |  | yes |
| Heparin |  |  |  |  |  |  | yes |
| Heparin |  | yes |  |  | yes |  |  |
| Heparin | yes |  |  | yes |  |  |  |
| Heparin | yes |  |  | yes |  |  |  |

### After controlling for potential confounding variables, patients that were administered enoxaparin have lower mortality rates

To simultaneously account for the effects of a range of possible confounders, we also examined statistical differences in outcomes between 1:1 propensity matched cohorts of patients who received enoxaparin but not heparin vs patients who received heparin but not enoxaparin. These cohorts were originally 464 and 207 patients, respectively; after 1:1 propensity score matching we were left with 2 cohorts of size 143; we refer to these cohorts as the “matched enoxaparin” and “matched heparin” cohorts respectively. Quality of balance between covariates is shown in **Table 1**. Most covariates (including demographics, comorbidities, and conditions on admission) are well-matched.

Of the 158 patients in the matched enoxaparin cohort, 8 (5.6%) were eventually deceased; in the matched heparin cohort, 21 (15%) were deceased (**Table 3**). The rate of mortality in the matched heparin cohort was higher (chi-square p-value 0.02; adjusted p-value 0.09) (**Table 3**). The risk ratio of mortality for heparin patients is 2.63 (95% CI: [1.18, 5.41]). Mean hospital length of stay (among alive patients) for matched heparin was 6.9 days vs 5.8 days for matched enoxaparin. Mean ICU length of stay (among alive patients) for matched heparin was 1.9 days vs 1.1 days for matched enoxaparin patients. However, neither difference was significant (hospital duration adjusted p-value: 0.17; ICU duration adjusted p-value 0.17).

Difference in rates of certain complications among the matched cohorts were analyzed. Complications that occurred at a higher rate in the matched heparin cohort than the matched enoxaparin cohort include acute kidney injury (7.7% vs 1.4%, chi-square p-value 0.2, BH-adjusted p-value 0.10) and bacterial pneumonia (5.6% vs 0%, chi-square p-value 0.01, BH-adjusted p-value 0.10) (**Table 6**). However, we note that due to small cohort sizes none of these differences were statistically significant after taking into account the adjustment for multiple hypotheses.

**Table 6:** Summary of occurrences of complications during hospitalization (days 0 to 28) for **matched** cohorts of hospitalized COVID-19 patients who have taken either heparin or enoxaparin.

| Complication | Enoxaparin, but not Heparin (Matched) | Heparin, but not Enoxaparin (Matched) | Chi-square p-value | BH-adjusted p-value |
| --- | --- | --- | --- | --- |
| Number of patients | 143 | 143 |  |  |
| Acute kidney injury | 2 (1.4%) | 11 (7.7%) | 0.02 | 0.10 |
| Bacterial pneumonia | 0 (0%) | 8 (5.6%) | 0.01 | 0.10 |
| Cardiac arrest | 0 (0%) | 0 (0%) | 1.00 | 1.00 |
| Cardiac arrhythmia | 2 (1.4%) | 1 (0.7%) | 1.00 | 1.00 |
| Co- or secondary infection | 1 (0.7%) | 7 (4.9%) | 0.07 | 0.22 |
| Liver dysfunction | 3 (2.1%) | 2 (1.4%) | 1.00 | 1.00 |
| Pleural Effusions | 0 (0%) | 0 (0%) | 1.00 | 1.00 |
| Stroke / Cerebrovascular incidents | 3 (2.1%) | 1 (0.7%) | 0.61 | 1.00 |
| Viral pneumonitis | 24 (17%) | 20 (14%) | 0.62 | 1.00 |

### Augmented curation of EHR patient notes shows that patients that were administered enoxaparin have lower rates of thrombotic events

We aimed to see if there were differences in occurrence of thrombotic events amongst patients treated strictly with Heparin and those treated strictly with Enoxaparin. To do so, we used a BERT-based neural network to extract thrombotic events from EHR notes and classify the sentences sentiment. If the note was dated within the timeframe of up to 28-days after that patient’s first hospitalization date and contained positive sentiment for a thrombotic event (with >= 0.9 confidence), a patient was considered to have experienced that thrombotic event. Similarly, we identified patients that have thrombotic comorbidities. Patients were considered as having a thrombotic comorbidity if they experienced a thrombotic event up to a year prior to their first hospitalization date. Thrombotic events considered were deep vein thrombosis, pulmonary embolism, myocardial infarction, venous thromboembolism, thrombotic stroke, cerebral venous thrombosis, and disseminated intravascular coagulation.

We found that, across all thrombotic phenotypes, there is a greater or equal rate of complications during the study period (days 0 to 28) for the Heparin cohort relative to the Enoxaparin cohort (risk ratio: 5.7, 95% C.I.: [2.1, 33.9], p-value: 0.024). In particular, for deep vein thrombosis, the rate is 0.2% in the enoxaparin cohort versus 2.9% in the heparin cohort (risk ratio: 13.5, 95% C.I.:[1.7, 56.8], p-value: 0.01, adjusted p-value: 0.04) (**Table 7**). Note that in the year leading up to the study period, there is not a statistically significant difference between these two cohorts (risk ratio: 1.5, 95% C.I.:[0.47, 5.1], p-value: 0.77, adjusted p-value: 1.0) (**Table 8**).

**Table 7:** Comparison of thrombotic events from Day 0 to Day 28. Counts of patients with thrombotic events reported in the clinical notes in days 0 to 28 among the cohorts administered Heparin only and Enoxaparin only, along with p-values from Chi-squared statistical tests. Presence of thrombotic events in the clinical notes was determined via a BERT-based neural network model. The thrombotic events considered included: deep vein thrombosis (DVT), cerebral venous thrombosis (CVT), venous thromboembolism (VTE), disseminated intravascular coagulation (DIC), pulmonary embolism (PE), myocardial infarction (MI), and cerebral stroke.

| **Phenotype** | **Enoxaparin, but not Heparin (Original)** | **Heparin, but not Enoxaparin (Original)** | **Chi-square p-value** | **BH-adjusted p-value** |
| --- | --- | --- | --- | --- |
| Deep vein thrombosis (DVT) | 1 (0.2%) | 6 (2.9%) | 0.01 | 0.04 |
| Cerebral venous thrombosis (CVT) | 0 (0.0%) | 0 (0.0%) | 1.00 | 1.00 |
| Venous thromboembolism (VTE) | 0 (0.0%) | 1 (0.5%) | 0.68 | 1.00 |
| Disseminated intravascular coagulation (DIC) | 0 (0.0%) | 0 (0.0%) | 1.00 | 1.00 |
| Pulmonary embolism (PE) | 1 (0.2%) | 3 (1.4%) | 0.17 | 0.41 |
| Myocardial infarction (MI) | 0 (0.0%) | 2 (1.0%) | 0.18 | 0.41 |
| Cerebral stroke | 0 (0.0%) | 0 (0.0%) | 1.00 | 1.00 |
| Total patients with at least 1 thrombotic event | 2 (0.4%) | 9 (4.3%) | 7.8E-04 |  |
| Total patients | 464 | 207 |  |  |

**Table 8:** Comparison of thrombotic events from Day −365 to Day −1. Counts of patients with thrombotic events reported in the clinical notes in days −365 to −1 among the cohorts administered Heparin only and Enoxaparin only, along with p-values from Chi-squared statistical tests. Presence of thrombotic events in the clinical notes was determined via a BERT-based neural network model. The thrombotic event phenotypes considered included: deep vein thrombosis (DVT), cerebral venous thrombosis (CVT), venous thromboembolism (VTE), disseminated intravascular coagulation (DIC), pulmonary embolism (PE), myocardial infarction (MI), and cerebral stroke.

| **Phenotype** | **Enoxaparin, but not Heparin (Original)** | **Heparin, but not Enoxaparin (Original)** | **Chi-square p-value** | **BH-adjusted p-value** |
| --- | --- | --- | --- | --- |
| Deep vein thrombosis (DVT) | 6 (1.3%) | 4 (1.9%) | 0.77 | 1.00 |
| Cerebral venous thrombosis (CVT) | 0 (0.0%) | 0 (0.0%) | 1.00 | 1.00 |
| Venous thromboembolism (VTE) | 0 (0.0%) | 0 (0.0%) | 1.00 | 1.00 |
| Disseminated intravascular coagulation (DIC) | 0 (0.0%) | 0 (0.0%) | 1.00 | 1.00 |
| Pulmonary embolism (PE) | 4 (0.9%) | 2 (1.0%) | 0.76 | 1.00 |
| Myocardial infarction (MI) | 3 (0.6%) | 7 (3.4%) | 0.02 | 0.13 |
| Cerebral stroke | 1 (0.2%) | 1 (0.5%) | 0.86 | 1.00 |
| Total patients with at least 1 thrombotic event | 12 (2.6%) | 12 (5.8%) | 0.07 |  |
| Total patients | 464 | 207 |  |  |

## Discussion

This retrospective study highlights interesting differences in the outcomes associated with anticoagulants in COVID-19 patients, warranting further follow up. A few caveats are in order. Compounding this limitation, enoxaparin and heparin differ in FDA label indications. The label for enoxaparin^13^ includes the prophylaxis and treatment of deep vein thrombosis (DVT) with or without pulmonary embolism (PE) in various settings, the prophylaxis of ischemic complications of unstable angina and non-Q-wave myocardial infarction (MI), and the treatment of acute ST-segment elevation MI managed medically or with subsequent percutaneous coronary intervention. The label for heparin^14^ includes includes similar prophylactic indications as well as the treatment of a broader spectrum of acute embolic events including peripheral arterial embolism and embolism in the setting of atrial fibrillation, the treatment of consumptive coagulopathies, and usage as an anticoagulant in high-risk patient groups such as those undergoing blood transfusions, extracorporeal circulation, and dialysis procedures. Furthermore, it is important to analyze whether these medications were administered as prophylactics or in treatment for a complication. Thus, it is possible that the patient population receiving heparin is more severely or acutely ill to begin with. In follow-up analyses, it may be interesting to consider initial laboratory values such as eGFR as a measure of baseline liver function as well.

Nevertheless, accumulating evidence suggests that enoxaparin may be more efficacious than heparin in certain cases such as for the treatment of acute coronary syndromes^15^. Also of interest as a driver of differential outcomes is the phenomenon of Heparin Induced Thrombocytopenia (HIT), a drug reaction associated with decreased platelet count and a high risk of thrombosis. HIT is caused by platelet-activating antibodies against PF4/heparin complexes and has been shown to occur in some circumstances in COVID-19 patients treated with unfractionated heparin^16^. These findings highlight the plausibility of a biological underpinning to the observations reported in the present work. Overall, these results motivate future studies and trials that could enable the development of more efficacious standard of practice in regards to administration of anticoagulants in COVID-19 patients.

## Methods

### Study Design

This research was conducted under IRB 20-003278, “Study of COVID-19 patient characteristics with augmented curation of Electronic Health Records (EHR) to inform strategic and operational decisions”, approved by the Mayo Clinic’s institutional review board (IRB). Under this IRB, an observational study was conducted with a study population of 1108 patients, defined as hospitalized patients who tested positive for SARS-CoV-2 within 7 days prior to admission or during admission to the hospital. Data available for analysis includes patient demographics (age, gender, race, etc.) as well as clinical data (medications administered, vital signs, ICD-10 diagnoses, etc.). Using this information, a dataset was assembled including key covariates and outcomes measures derived from structured clinical data. Covariates of interest include age, height, weight, comorbidities, admissions diagnoses, oxygenation methods, etc.

ICD-10 diagnosis data was consumed and used to define three sets of diagnoses of interest for the patient population. Complications are defined as diagnoses recorded during the patient’s treatment process. Admission diagnoses are defined as diagnoses made on the day of admission to the hospital, or within 7 day prior to admission. Comorbidities are defined as diagnoses made from a year prior to admission up to 7 days prior to admission to the hospital. For each of these categories a number of conditions were specified as being of interest and ICD-10 codes were selected to represent each of these conditions. ICD-10 codes used in this analysis are included in the supplement (see **Supplementary Material**).

To compare the outcomes of patients taking different anticoagulants, two cohorts were constructed: (i) patients who were administered enoxaparin but not heparin and (ii) patients who were administered heparin but not enoxaparin. The cohort sizes were 464 and 207 respectively. A number of statistical tests were conducted to identify any differences in outcomes. Statistical tests were applied to a number of outcomes (with Benjamini-Hochberg procedure applied to account for the problem of multiple comparisons; details below). Initially the following 5 outcomes were analyzed: (1) mortality status (i.e. was the patient ever recorded as deceased), (2) 28-day confirmed mortality status (i.e. of patients for whom we have have some confirmation of mortality status at 28 days following hospitalization, was the patient among the deceased), (3) ICU admission during hospitalization, (4) length of stay in the hospital (among alive patients), (5) length of stay in ICU (among alive patients).

To further characterize the outcomes of the 2 cohorts, occurrence of each of the following 9 complications from day 0 to day 28 were also analyzed: (1) acute kidney injury, (2) bacterial pneumonia, (3) cardiac arrest, (4) cardiac arrhythmia, (5) co- or secondary infection, (6) liver dysfunction, (7) pleural effusions, (8) stroke/cerebrovascular incident, and (9) Viral pneumonitis.

To account for potentially confounding variables, we performed propensity score matching to balance covariates between the two cohorts. The statistical tests for differences in outcomes were repeated on the matched cohorts. The covariates which were balanced include demographics, comorbidities and various features on admission. Further detail on the procedure, including a listing of covariates used, is below.

### Risk Ratio

Risk Ratios for Admission to the ICU and Mortality Status were calculated for patient cohorts defined by anticoagulant usage and presence of select comorbidities. Risk Ratio across two cohorts of interest is calculated by dividing the proportion of cohort 1 which responds affirmatively to a feature (or all members of a set of features) by the proportion of cohort 2 which responds affirmatively to the same feature (or set of features). Risk Ratio along with accompanying 95% confidence intervals for each cohort pair are calculated.

### Chi-Square

A Pearson’s Chi-Square test was run to test for statistically significant differences in proportion of occurrence of features across cohort pairs. This was done using the stats.chi2_contingency function of the scipy python package. Test statistics and p-values are reported for each cohort pair.

### t-Test

A two-sided t-test was performed to identify statistically significant differences in Hospital Length of Stay and ICU Length of Stay across two cohorts of patients who were discharged from the hospital alive and received different anticoagulants (enoxaparin and heparin). P-values are reported for these tests.

### Benjamini-Hochberg correction

As we are comparing multiple outcomes between two anticoagulant cohorts, a Benjamini-Hochberg correction is applied to a set of outcomes. Mortality was identified as a primary outcome prior to performing the tests, so it may not be necessary to apply the Benjamini-Hochberg correction on that p-value; however, adjusted p-values are reported for all outcomes.

### Propensity score matching

The two cohorts that were balanced were (i) the 464 patients who were administered enoxaparin but not heparin and (ii) the 207 patients who were administered heparin but not enoxaparin. Propensity scores were computed by fitting a logistic regression model to predict which of the 2 cohorts the patient was in, as a function of the covariates (listed further below). After computing propensity scores, 1:1 matching was done, using a heuristic caliper of 0.1 x pooled std deviation and allowing for drops.^17^ 143 matched pairs were found. After checking for quality of cohort balance (see **Table 1**), the same statistical procedures (chi-square test, t-test, risk ratio). were then run on the two matched cohorts of 143 patients each to identify if differences in outcome persist after the adjustments.

The covariates used for balancing are the following. Note all variables below except for age are binary.

- **Demographics**: Age, Gender, Race, Ethnicity
- **Comorbidities:** whether or not the patient has each of the following comorbidities, (1) cancer, (2) cardiac arrhythmias, (3) chronic kidney disease, (4) chronic pulmonary disease, (5) dementia, (6) depression, (7) diabetes, (8) hypertension, (9) obesity, (10) stroke or other neurological disorders
- **In ICU on admission to hospital**
- **Diagnosis on admission:** whether or not the patient is reported to have each of the following conditions on admission to hospital. (1) acute kidney injury, (2) bacterial pneumonia, (3) cardiac arrhythmias, (4) delirium / encephalopathy, (5) sepsis, (6) stroke
- **Oxygenation status on day of admission:** whether or not the patient is reported to have received the following forms of oxygen therapy on admission

### Augmented curation of thrombotic event phenotypes from the unstructured text of the Electronic Health Record (EHR) clinical notes

We used a previously developed state-of-the-art BERT-based neural network to classify sentiment of clinical manifestations and diagnoses in the EHR. Specifically, the model extracts sentences containing clinical phenotypes and classifies their sentiment into the following categories: Yes (confirmed clinical manifestation or diagnosis), No (ruled out clinical manifestation or diagnosis), Maybe (possibility of clinical manifestation or diagnosis), and Other (alternate context, e.g. family history of disease). The model was trained using 18,490 sentences and approximately 250 phenotypes with an emphasis on cardiovascular, pulmonary, and metabolic phenotypes. It achieves 93.6% overall accuracy and over 95% precision and recall for both “Yes” and “No” sentiment classification.

## Data Availability

All relevant insights are self-contained in the manuscript file and the underlying data was accessed under an IRB approved by the Mayo Clinic

## Supplementary Material

**Supplementary Table S1.**
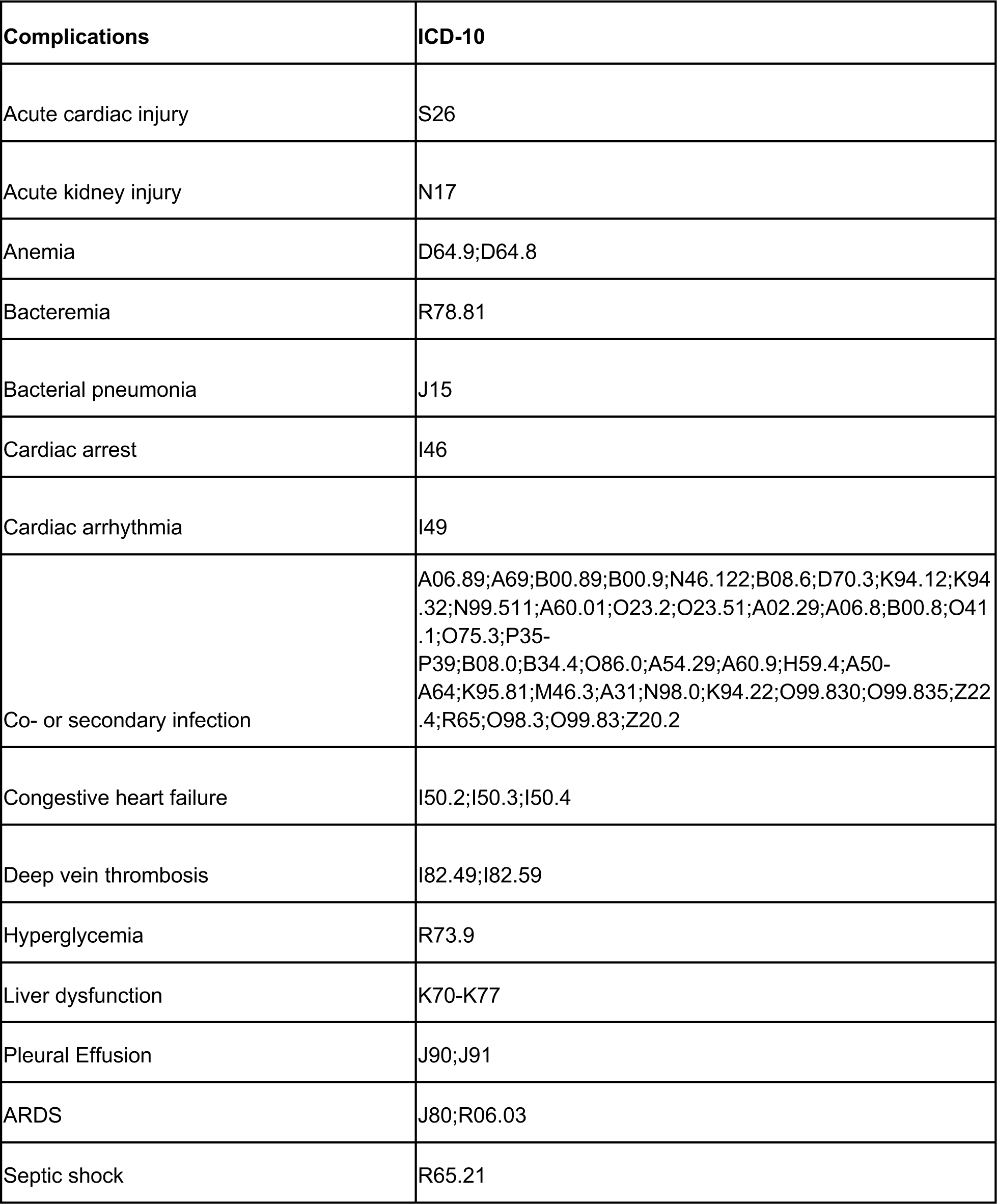
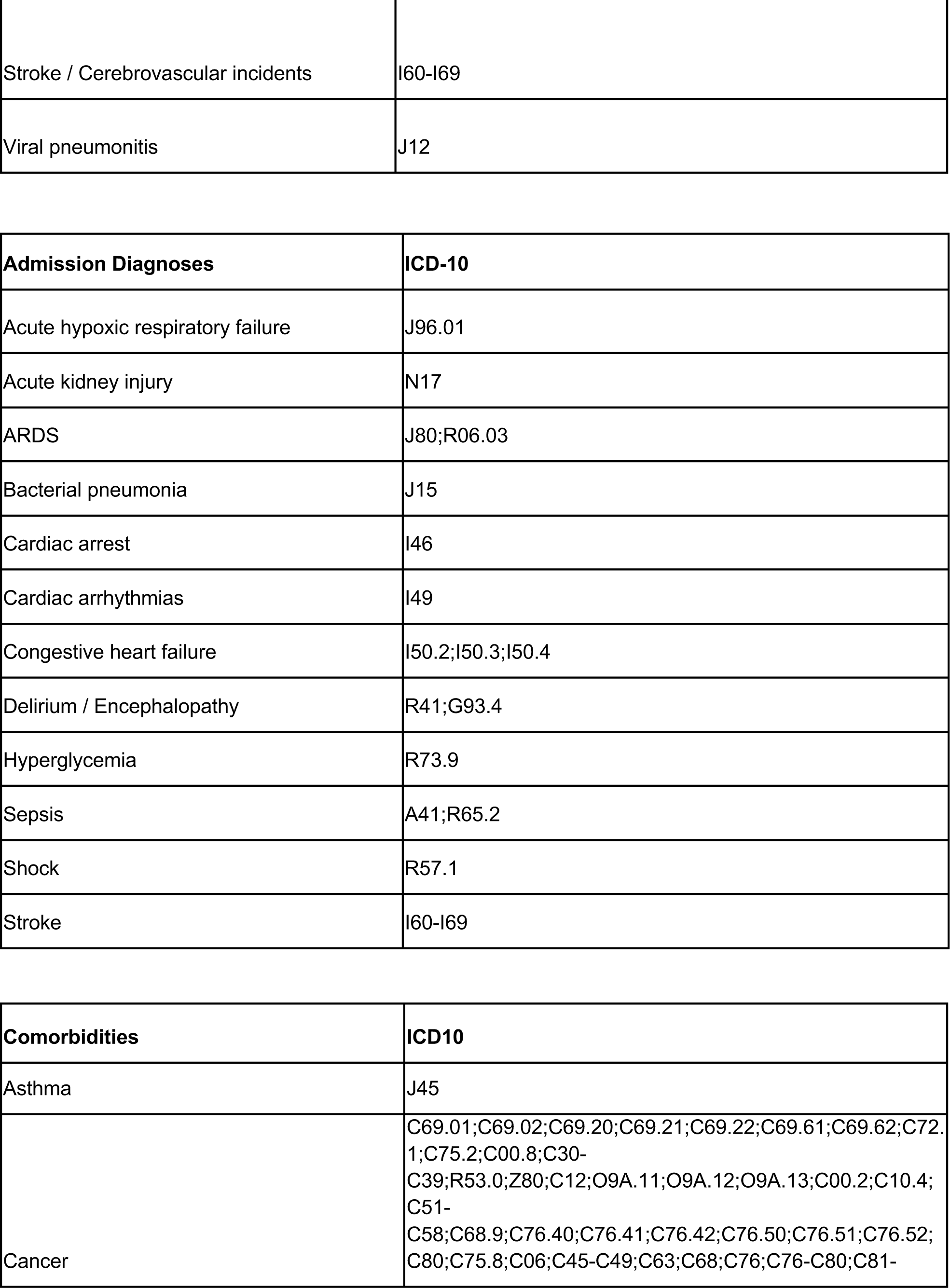
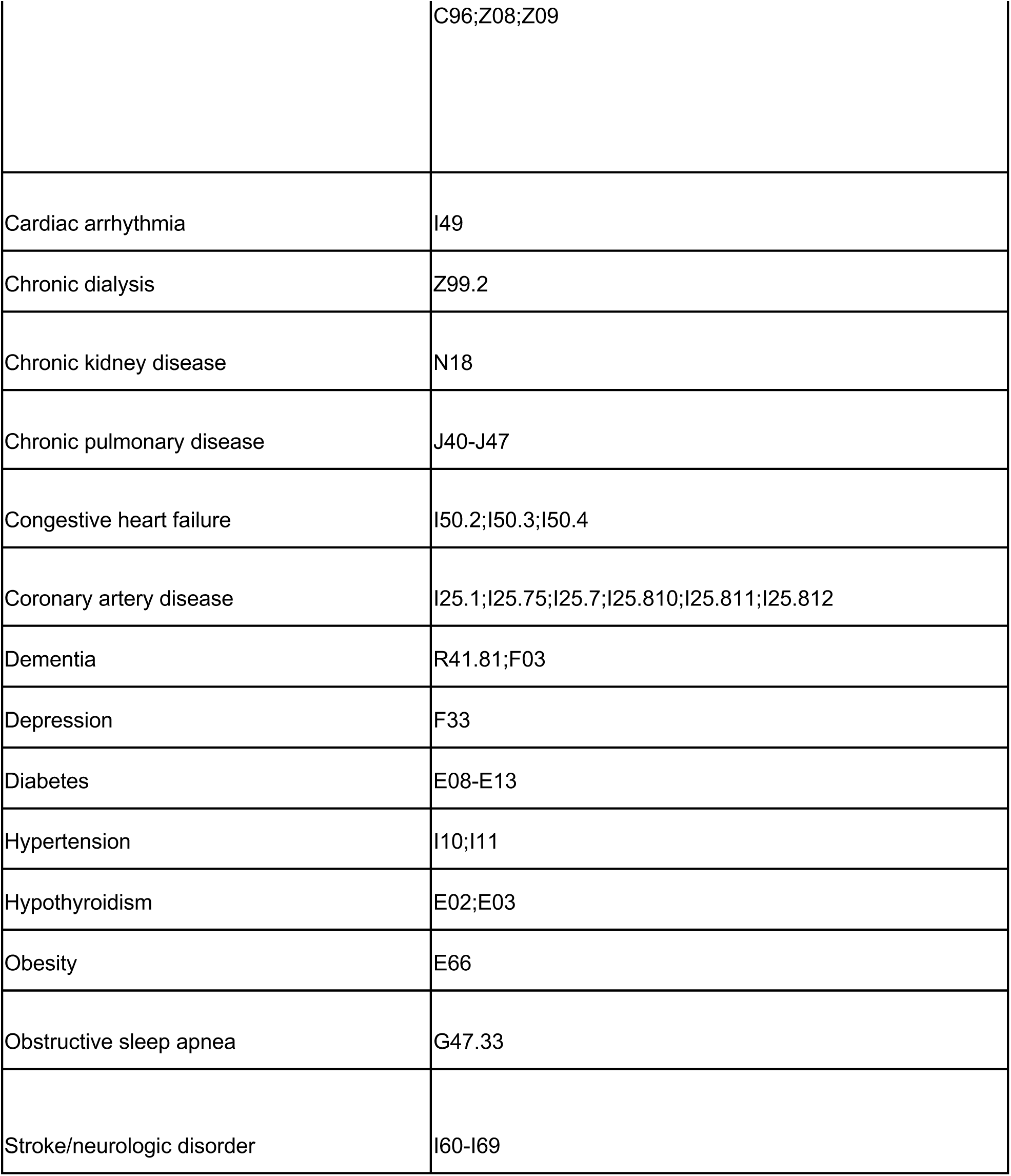
Diagnosis Code Definitions.

## References

1. Gandhi, R. T., Lynch, J. B. & del Rio, C. Mild or Moderate Covid-19. New England Journal of Medicine (2020) doi:10.1056/nejmcp2009249.

2. Wagner, T. et al. Augmented curation of clinical notes from a massive EHR system reveals symptoms of impending COVID-19 diagnosis. Elife 9, (2020).

3. Richardson, S. et al. Presenting Characteristics, Comorbidities, and Outcomes Among 5700 Patients Hospitalized With COVID-19 in the New York City Area. JAMA 323, 2052–2059 (2020).

4. Thachil, J. et al. ISTH interim guidance on recognition and management of coagulopathy in COVID-19. Journal of Thrombosis and Haemostasis vol. 18 1023–1026 (2020).

5. Klok, F. A. et al. Incidence of thrombotic complications in critically ill ICU patients with COVID-19. Thromb. Res. 191, 145–147 (2020).

6. Pawlowski, C. et al. Inference from longitudinal laboratory tests characterizes temporal evolution of COVID-19-associated coagulopathy (CAC). Elife 9, (2020).

7. Assistant Secretary for Public Affairs (ASPA). Fact Sheet: Explaining Operation Warp Speed. https://www.hhs.gov/coronavirus/explaining-operation-warp-speed/index.html (2020).

8. Connors, J. M. & Levy, J. H. COVID-19 and its implications for thrombosis and anticoagulation. Blood 135, 2033–2040 (2020).

9. Tang, N. et al. Anticoagulant treatment is associated with decreased mortality in severe coronavirus disease 2019 patients with coagulopathy. J. Thromb. Haemost. 18, 1094–1099 (2020).

10. Barco, S. et al. Enoxaparin for primary thromboprophylaxis in ambulatory patients with coronavirus disease-2019 (the OVID study): a structured summary of a study protocol for a randomized controlled trial. Trials 21, 770 (2020).

11. Tandon, R. et al. Effective Inhibition of SARS-CoV-2 Entry by Heparin and Enoxaparin Derivatives. bioRxiv (2020) doi:10.1101/2020.06.08.140236.

12. Lemos, A. C. B. et al. Therapeutic versus prophylactic anticoagulation for severe COVID-19: A randomized phase II clinical trial (HESACOVID). Thromb. Res. 196, (2020).

13. [No title]. https://www.accessdata.fda.gov/drugsatfda_docs/label/2009/020164s085lbl.pdf.

14. [No title]. https://www.accessdata.fda.gov/drugsatfda_docs/label/2017/017029s140lbl.pdf.

15. Murphy, S. A. et al. Efficacy and safety of the low-molecular weight heparin enoxaparin compared with unfractionated heparin across the acute coronary syndrome spectrum: a meta-analysis. Eur. Heart J. 28, 2077–2086 (2007).

16. Daviet, F. et al. Heparin Induced Thrombocytopenia in Severe COVID-19 Patients. Circulation (2020) doi:10.1161/CIRCULATIONAHA.120.049015.

17. Wang, Y. et al. Optimal Caliper Width for Propensity Score Matching of Three Treatment Groups: A Monte Carlo Study. PLoS One 8, e81045 (2013).

